# Greater lean-body-mass decline with tirzepatide than semaglutide in routine care, revealed by body-composition digital phenotyping

**DOI:** 10.64898/2026.04.11.26350687

**Authors:** Karthik Murugadoss, A.J. Venkatakrishnan, Venky Soundararajan

**Affiliations:** nference, 1 Main Street, East Arcade building, Cambridge, Massachusetts, USA; Metabolism Agentic Intelligence Atlas (MAIA), Cambridge, Massachusetts, USA

**Author notes:** Correspondence: Venky Soundararajan.

## Abstract

GLP-1 receptor agonists induce substantial weight loss, but the extent to which lean tissue and physical function are preserved in routine care remains poorly understood. Using an EHR-linked body-composition digital phenotyping pipeline with LLM-based extraction, we performed a large-scale longitudinal analysis of 670,422 first-episode GLP-1RA users, including 456,742 treated with semaglutide and 213,680 treated with tirzepatide. Among these, 7,965 individuals with paired pre- and post-initiation body-composition measurements were analyzed over 12 months. Tirzepatide was associated with greater relative lean body mass (LBM) loss than semaglutide at each measured time point, with excess LBM losses of 1.1%, 1.5%, 1.3% and 2% at 3, 6, 9 and 12 months, respectively. A *Depletive GLP-1 metabotype*, defined as >20% total body weight (TBW) loss with >5% LBM loss, was significantly more frequent with tirzepatide than semaglutide during the first year of therapy (10.3% versus 6.7%, p<0.001). By contrast, a *Prime GLP-1 metabotype*, defined as >10% TBW loss with <5% LBM loss, was numerically more frequent with semaglutide than tirzepatide, but not significantly so (12.3% versus 11.8%, p=0.66). Higher drug dose and longer exposure were associated with progressively greater LBM decline in both treatment groups (both p<0.001). Among 3,746 examined EHR phenotypes, baseline musculoskeletal pain emerged as the most significant correlate of greater LBM loss (BH-adjusted q<0.001): cervicalgia (semaglutide, −4.1 percentage points; tirzepatide, −14.3 percentage points) and knee pain (semaglutide, −4.8 percentage points; tirzepatide, −13.4 percentage points), consistent with mobility-limited patients being more vulnerable to lean-tissue depletion during incretin therapy. Analysis of EHR notes for on-treatment functional features showed reduced exercise tolerance was the strongest correlate of greater LBM loss, increasing by 7.2 and 11.1 percentage points in semaglutide- and tirzepatide-treated patients, respectively. An independent analysis of all available Single-cell RNA-seq data from human musculature showed broader GIPR+ cellular distribution than GLP1R+ cells across immune, stromal, vascular, and contractile compartments, providing plausible biological context for the greater LBM loss observed in routine care with tirzepatide (dual GLP1R-GIPR agonist) relative to semaglutide (GLP1R-specific agonist). In this observational study, greater weight-loss efficacy did not necessarily translate into more favorable body-composition outcomes, underscoring the need for clinical decision-making and trial designs that maximize each patient’s likelihood of achieving a *Prime GLP-1 metabotype*.

## Introduction

Semaglutide and tirzepatide have reshaped pharmacologic management of obesity. Randomized trials, observational comparisons and the first head-to-head obesity trial consistently show greater mean total-weight reduction with tirzepatide than with semaglutide^1–7^. Body weight loss alone, however, cannot distinguish between adipose loss and attrition of muscle-containing lean tissue, a distinction that matters for metabolic health, physical function, broader quality of life for patients, and their long-term adherence to as well as durability of benefit^8–13^. Prior body-composition studies indicate that both agents reduce fat mass substantially while also lowering lean mass^8,14– 18^. Exploratory analyses from STEP 1 and related semaglutide programs showed mixed-tissue weight loss rather than selective adipose reduction alone^19^. SURMOUNT-1 body-composition analyses reported the same overall pattern for tirzepatide^15^. At the same time, recent reviews emphasize that lean body mass is not synonymous with skeletal muscle mass, that relative lean proportion often increases during incretin-based weight loss, and that functional outcomes may be more informative than absolute lean-mass change alone^9,20^.

A central gap is that most body-composition data arise from protocolized trials or small mechanistic cohorts, whereas real-world patients differ in baseline phenotype, measurement cadence, treatment persistence and ascertainment modality^2,8^. Real-world comparative studies of semaglutide and tirzepatide have largely focused on scale weight rather than tissue composition, leaving uncertainty about whether the superior weight-loss efficacy of tirzepatide is accompanied by systematically different lean-tissue trajectories^8,9^.

Mechanistically, tirzepatide’s additional engagement of the glucose-dependent insulinotropic polypeptide receptor (GIPR) has been proposed to shape not only appetite and glycemic physiology but also tissue remodeling programs that could, in principle, influence lean-tissue dynamics^17,21^. However, the mechanism through which GIPR might influence physiology in muscle-resident cell types remains incompletely understood, motivating receptor-topology approaches using public single-cell atlases to contextualize clinical observations^22^.

Here, we integrated longitudinal EHR-derived paired body-composition measurements of total body weight (TBW) and lean body mass (LBM) with phenotype stratification and functional outcome analyses to compare real-world trajectories of total body weight, fat mass and lean body mass after semaglutide or tirzepatide initiation, incorporating clinical records of GLP-1RA adherence. Among patients with documented baseline and follow-up body-composition measurements and evidence of on-treatment exposure, we quantified both absolute and relative compositional change, asked whether deeper weight loss was accompanied by differential lean-tissue loss across clinically distinct subgroups, and linked these patterns to post-treatment functional correlates derived from procedure codes and natural-language processing of clinical notes. Further, we used single-cell GLP1R and GIPR expression maps from human musculature associated datasets to place the observed between-drug clinical differences in biological context. This design moves beyond prior trial-based and head-to-head weight-loss comparisons by combining real-world paired body-composition trajectories with phenotype stratification and functional readouts.

## Results

### Real-world metabolic deep phenotyping available for a fraction of new GLP-1 users

Across 670,422 first-episode GLP-1 patients, 456,742 initiated semaglutide and 213,680 initiated tirzepatide (**Fig. 1A**). Of these, 7,965 patients had lean body mass (LBM) measurements before (baseline) and after initiation of their first prescription of semaglutide (N=6,196) or tirzepatide (N=1,769). Detailed demographics can be found in **Table 1**. The LBM measurements were obtained using various body composition measurement techniques including Bioelectric Impedance Analysis (BIA), DEXA, or Bod Pod (Air Displacement Plethysmography), with almost 85% of the study population undergoing serial BIA.

**Table 1:** Baseline demographic, clinical, and treatment characteristics of the semaglutide and tirzepatide cohorts included in the body-composition analysis. Values are presented as mean (SD), median [IQR], or percentage, as appropriate.

| Characteristic | Semaglutide (n=6,196) | Tirzepatide (n=1,769) |
| --- | --- | --- |
| **Demographics** |  |  |
| Age at index, (years) | 46.2 (15.0) | 48.5 (14.0) |
| Female, % | 78.50% | 74.60% |
| **Baseline Characteristics** |  |  |
| T2DM, % | 11.60% | 10.50% |
| Baseline BMI, kg/m<sup>2</sup> | 38.1 (4.5) | 37.4 (4.5) |
| Baseline weight, kg | 113.4 (24.2) | 111.8 (24.2) |
| Baseline lean body mass, kg | 56.9 (15.1) | 57.4 (16.0) |
| Prior GLP-1RA exposure, % | 13.30% | 9.00% |
| **Treatment characteristics** |  |  |
| Treatment duration, days | 381 [200, 705] | 258 [180, 433] |
| Number of prescriptions | 6 [2, 10] | 5 [2, 9] |

**Fig. 1.**
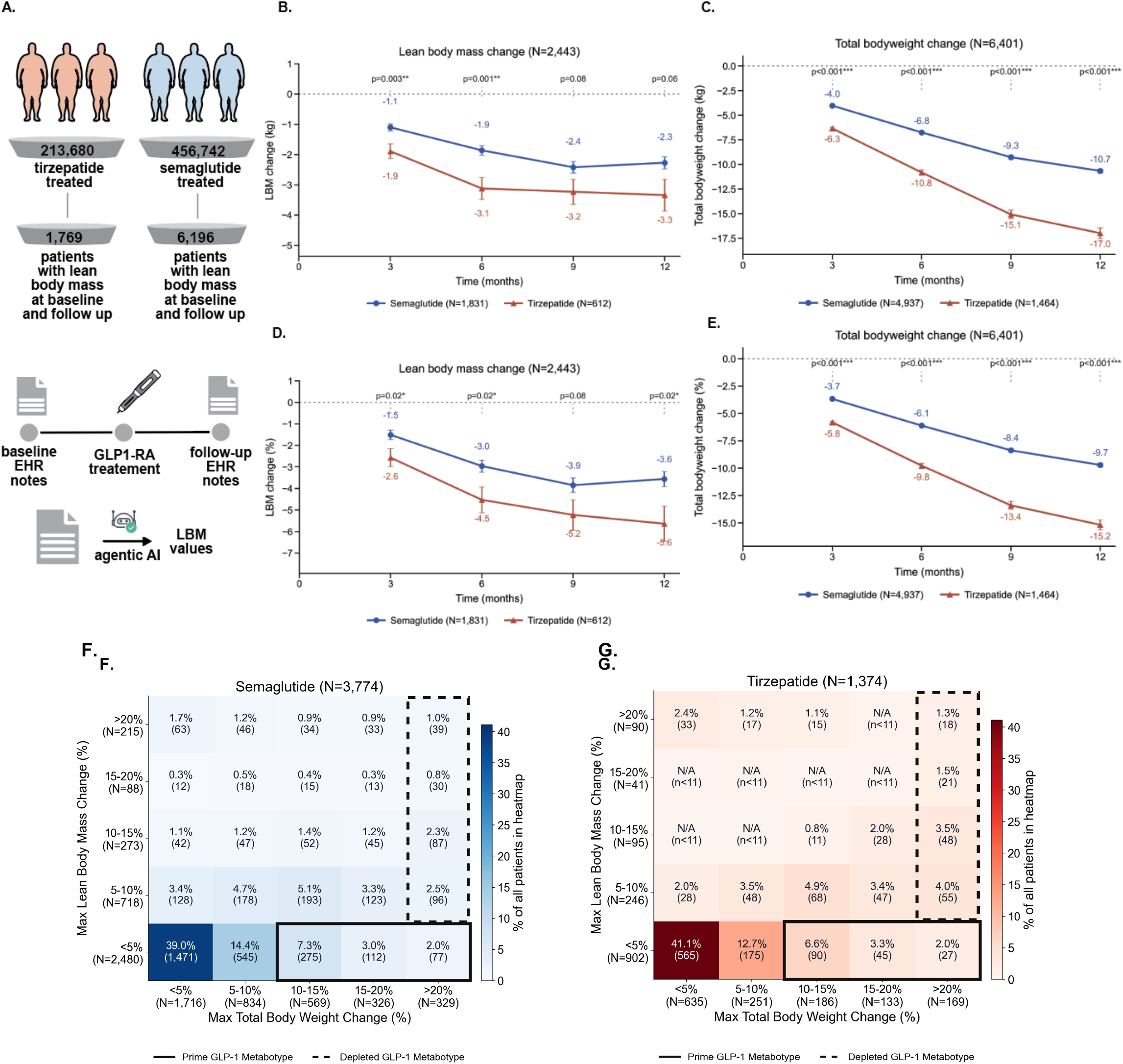
Total body weight (TBW) loss and lean body mass (LBM) loss response of patients on semaglutide or tirzepatide. **A**. Cohort selection funnel used to query the nSights federated EHR network of 29 million patient lives, specifically selecting patients with first GLP-1RA exposure on semaglutide or tirzepatide that contain a baseline body composition measurement and at least one additional measurement during the following 12 months on GLP-1RA. **B**. Absolute lean body mass response (in kg) across 12 months on semaglutide patients (blue) and tirzepatide (red). **C**. As in B, but reporting absolute total body weight response (in kg). **D**. As in B, but reporting % lean body mass change from baseline at t = 0 mo. **E**. As in B, but reporting % total body weight change from baseline at t = 0 mo. **F**. Heatmap of semaglutide patient response stratified by maximum % total body weight change (x-axis: <5%, 5–10%, 10–15%, 15–20%, >20% loss) and maximum % lean body mass change (y-axis: <5%, 5–10%, 10–15%, 15–20%, >20% loss). For each tile, the percentage and count of patients meeting those loss criteria are reported; tile color reflects prevalence relative to the overall cohort. The solid box denotes the *Prime GLP-1 Metabotype*, defined as ≥10% total body weight loss with <5% lean body mass loss, capturing patients with pronounced fat-preferential weight reduction. The dashed box denotes the *Depletive GLP-1 Metabotype*, defined as >20% total body weight loss with ≥5% lean body mass loss, capturing patients with clinically significant lean mass depletion alongside maximal weight loss. **G**. As in F, but for the tirzepatide cohort.

### Tirzepatide was consistently associated with greater LBM loss than semaglutide over a 12 month period

Tirzepatide showed greater absolute LBM loss than semaglutide at every measured time point (**Fig. 1B**). Mean absolute LBM change with semaglutide was −1.1 kg (s.d. 5.0) at 3 months, −1.9 kg (s.d. 5.4) at 6 months, −2.4 kg (s.d. 5.5) at 9 months and −2.3 kg (s.d. 6.0) at 12 months, compared with −1.9 kg (s.d. 6.0), −3.1 kg (s.d. 6.4), −3.2 kg (s.d. 5.6) and −3.3 kg (s.d. 6.3), respectively, with tirzepatide. The between-drug difference therefore measured 0.8 kg at 3 months (p=0.003), 1.2 kg at 6 months (p=0.001), 0.8 kg at 9 months (p=0.08) and 1.0 kg at 12 months (p=0.06). The same pattern was observed on the relative scale, where LBM change was expressed as a percentage of each patient’s baseline lean body mass (**Fig. 1D**). Percentage LBM change from baseline with semaglutide was −1.5% (standard deviation 9.1), −3.0% (9.9), −3.9% (9.7) and −3.6% (10.4) at 3, 6, 9 and 12 months, respectively, compared with −2.6% (10.3), −4.5% (10.7), −5.2% (9.6) and −5.6% (9.8) with tirzepatide. Thus, tirzepatide was associated with greater relative LBM loss than semaglutide at every measured time point, with excess losses of 1.1% (p=0.02), 1.5% (p=0.02), 1.3% (p=0.08) and 2.0% (p=0.02) at 3, 6, 9 and 12 months, respectively.

Consistent with prior head-to-head clinical and real-world studies, absolute (**Fig. 1C**) and relative (**Fig. 1E**) total body weight (TBW) loss was greater with tirzepatide than with semaglutide across all measured time points. Mean bodyweight change with semaglutide was −3.7% (standard deviation 4.2), −6.1% (6.5), −8.4% (8.2) and −9.7% (9.6) at 3, 6, 9 and 12 months, respectively, compared with −5.8% (4.8), −9.8% (7.2), −13.4% (8.9) and −15.2% (10.1) with tirzepatide, yielding between-drug differences of 2.1% (p<0.001), 3.7% (p<0.001), 5.0% (p<0.001) and 5.5% (p<0.001), respectively. Thus, tirzepatide’s greater LBM loss occurred in the setting of greater total body weight reduction relative to semaglutide.

### Most patients in both treatment groups remained within low weight-loss and low LBM loss categories

In semaglutide-treated patients, 65.7% (n=2,480) of the full cohort fell in the <5% LBM-loss strata and 19.0% (n=718) fell in the 5 to 10% LBM-loss strata (**Fig. 1F**). The corresponding estimates in tirzepatide-treated patients were 65.6% (n=902) and 17.9% (n=246) (**Fig. 1G**). There was no significant difference in the proportion of patients in the low relative LBM loss (less than 5%) strata between semaglutide and tirzepatide (p=0.965).

On the other hand, greater relative LBM loss was far less common and occurred at similar rates for semaglutide and tirzepatide. For example, LBM reduction of >15% occurred in 8.0% (n=303) of semaglutide-treated patients and 9.5% (n=131) of tirzepatide-treated patients overall. Specifically, in semaglutide-treated patients, the 15-20% and >20% LBM-loss strata accounted for 2.3% (n=88) and 5.7% (n=215) of the cohort, respectively (**Fig. 1F**), compared to 3.0% (n=41, p=0.185) and 6.6% (n=90, p=0.251) of the tirzepatide cohort (**Fig. 1G**).

### Optimal outcome of high TBW loss coupled with low LBM loss (‘Prime GLP-1 Metabotype’) was achieved for a minority of patients taking semaglutide or tirzepatide

Seemingly minor reductions in LBM (>5% over 6-12 months) and the attendant reduction in strength lead to pathologies such as sarcopenia and render patients frail and at risk for falls and other adverse events^23^. Therefore, the most desirable body-composition outcome, defined here as the *Prime GLP-1 metabotype*, was low percentage LBM loss (<5%) combined with higher percentage TBW loss (>10%). The corresponding cells in the heatmaps in **Fig. 1F** (semaglutide) and **Fig. 1G** (tirzepatide) accounted for 12.3% (n=463) of semaglutide-treated patients and 11.8% (n=162) of tirzepatide-treated patients (p=0.66). For the Prime GLP-1 metabotype, the mean baseline BMI appeared significantly higher with semaglutide (38.0–38.7) than with tirzepatide (36.9–37.2) (p=0.011; **Fig. S1**). Together, these findings suggest that semaglutide conferred a modest but non-significant advantage over tirzepatide in the overall frequency of Prime GLP-1 metabotypes, whereas semaglutide-associated Prime GLP-1 metabotypes occurred at significantly higher baseline BMI.

### LBM loss remained a minority component of TBW loss, but clinically relevant depletion was still common

Although greater TBW loss was generally associated with upward movement into higher LBM-loss strata, high TBW loss did not rigidly map onto high LBM loss though they were similarly correlated. For example, 88.3% of semaglutide-treated patients (**Fig. 1F**) and 90.6% of tirzepatide-treated patients (**Fig. 1G**) showed equal or greater %TBW loss than %LBM loss, reinforcing that LBM loss is a minor component of overall weight loss on GLP-1RAs. However, regardless of TBW outcomes, 34.3% of semaglutide-treated patients and 34.3% of tirzepatide-treated patients demonstrated >5% LBM loss of muscle mass. Given the widespread usage of GLP-1RAs, large patient populations would benefit from superior LBM management.

### Depletive outcome of high TBW loss coupled with high LBM loss (‘Depletive GLP-1 Metabotype’) was more frequent with tirzepatide than semaglutide

At the same time, a Depletive GLP-1 metabotype, defined as >20% total body weight loss together with >5% lean body mass loss, was observed in 6.7% (n=252) of semaglutide-treated patients (**Fig. 1F**) and 10.3% (n=142) of tirzepatide-treated patients (**Fig. 1G**), indicating significant enrichment with tirzepatide relative to semaglutide (p<0.001). For the Depletive GLP-1 metabotype, baseline BMI appeared broadly comparable between semaglutide and tirzepatide (37.3–39.4 versus 36.6–39.9; p=0.37; **Fig. S1**).

By contrast, a more extreme unfavorable outcome, defined as ≥15% LBM loss together with ≥15% TBW loss, remained uncommon in both cohorts, accounting for only 3.0% (n=115) of the semaglutide cohort and 3.8% (n=52) of the tirzepatide cohort (p=0.187). Thus, although severe LBM depletion was not the dominant pattern even among patients with the largest TBW reductions, Depletive GLP-1 metabotypes were still significantly more frequent with tirzepatide than semaglutide, highlighting an important opportunity for future obesity therapeutics and clinical monitoring strategies to improve lean-tissue preservation.

### Higher drug exposure of both semaglutide and tirzepatide were strongly associated with progressively greater relative LBM loss

Both semaglutide and tirzepatide showed inverse associations between dose tier and percentage LBM change across the 12-month observation period, indicating progressively greater LBM loss at higher doses (**Fig. 2A-B**). For semaglutide, each 1 mg increase in dose was associated with an additional 1.9% decrease in LBM (R^2^ = 0.83) and an additional 3.95% decrease in TBW (R^2^ = 0.93) (**Fig. 2A, 2C**). For tirzepatide, each 1 mg increase in dose was associated with an additional 0.45% decrease in LBM (R^2^ = 0.87) and an additional 0.74% decrease in TBW (R^2^ = 0.82) (**Fig. 2B, 2D**). Despite these clear dose-dependent trends, there was marked within-dose-tier heterogeneity in both LBM and TBW response for both drugs, with wide interquartile ranges particularly evident at higher doses (**Fig. 2A-D**).

**Fig. 2.**
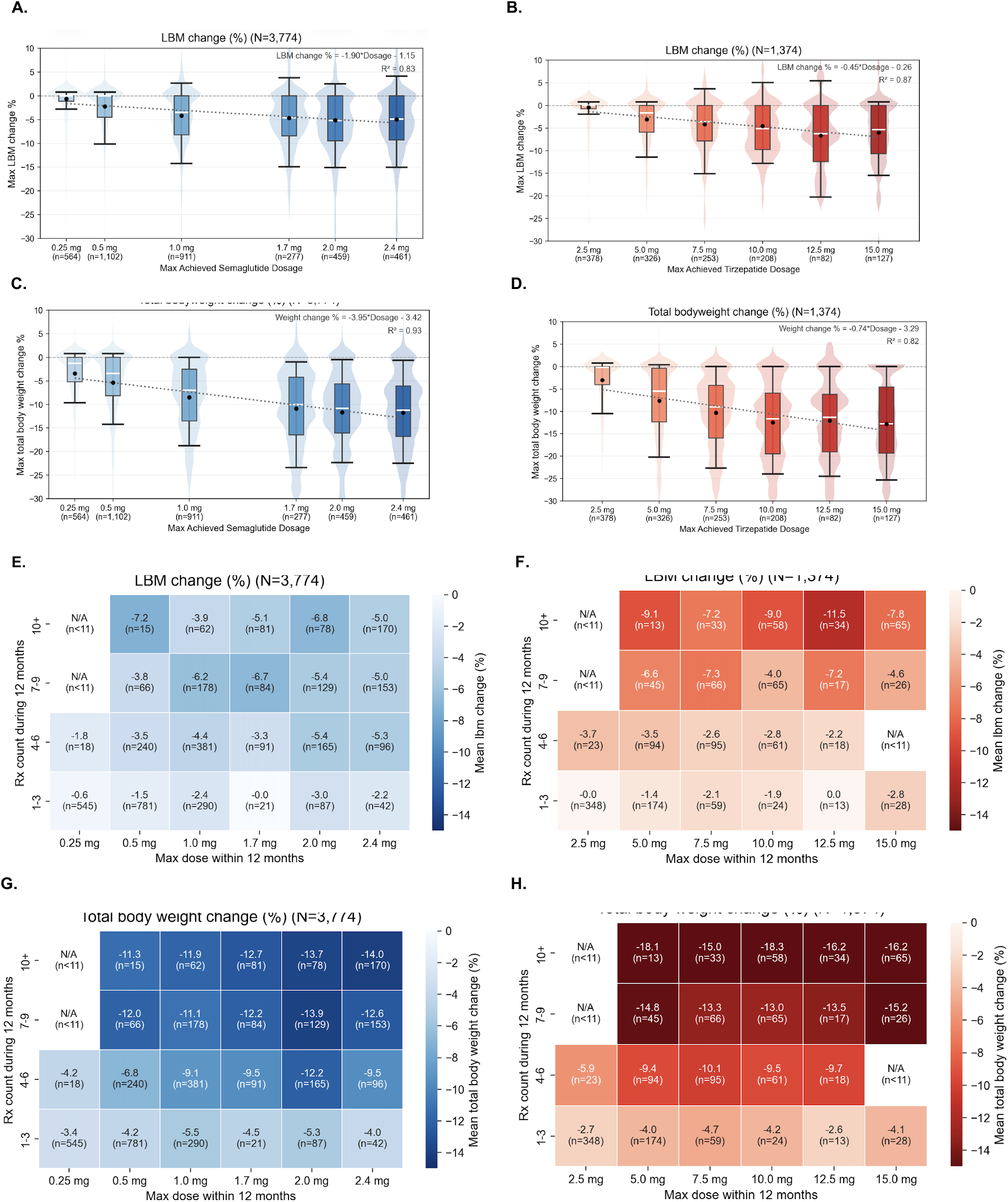
Drug exposure is correlated with total body weight loss and, more heterogeneously, to lean body mass loss. **A**. Box and whisker overlaid on violin plots of the relationship between % lean body mass change (y-axis) as a function of semaglutide dose on the x-axis. The black dot represents the mean. The white stripe represents the median. The box represents the IQR, whereas the bounds of the violin plot indicate the range. Linear regression through the median is represented by the black, dashed line. **B**. As in A, but for tirzepatide. **C**. As in A, but box and whisker overlaid on violin plots of the relationship between % total body weight change (y-axis) as a function of semaglutide dose on the y-axis. **D**. As in C, but for tirzepatide. **E**. Heatmap of semaglutide-treated patient % lean body mass change response as a function of semaglutide dose on the x-axis and number of semaglutide prescriptions during the 12 month follow-up period on the y-axis (stratified into 1-3 rx, 4-6 rx, 7-9 rx, and 10+ rx). For each tile, the % lean body mass change (and number of patients meeting these loss criteria) is reported and the tile is colored based on the magnitude of the % lean body mass change between the semaglutide and tirzepatide cohorts. **F**. As in E, but for tirzepatide. **G**. Heatmap of semaglutide-treated patient % total body weight change response as a function of semaglutide dose on the x-axis and number of semaglutide prescriptions during the 12 month follow-up period on the y-axis (stratified into 1-3 rx, 4-6 rx, 7-9 rx, and 10+ rx). For each tile, the % total body weight change (and number of patients meeting these loss criteria) is reported and the tile is colored based on the magnitude of the % total body weight change between the semaglutide and tirzepatide cohorts. **H**. As in G, but for tirzepatide.

Further, both drugs showed a clear association between total prescription exposure and total body weight loss, with progressively deeper TBW reduction across increasing prescription-count strata in semaglutide-treated (**Fig. 2G**) and tirzepatide-treated patients (**Fig. 2H**). In semaglutide-treated patients, TBW loss appeared to plateau between the 7-9 and 10+ prescription strata, although the difference in TBW change (1.0 percentage point) was not significant after accounting for dose (dose-adjusted model; p=0.209); in contrast, tirzepatide-treated patients continued to show deeper TBW loss at higher exposure levels, with dose-adjusted mean TBW change differing by an additional 3.04 percentage points between the 7-9 and 10+ prescription strata (p<0.001). LBM loss was more heterogeneous than TBW loss across dose and prescription strata for both semaglutide (**Fig. 2E**) and tirzepatide (**Fig. 2F**), although dose-by-exposure models still showed significant overall associations for both drugs (semaglutide: p<0.001, R^2^=0.088; tirzepatide: p<0.001, R^2^=0.194). In both treatment groups, greater prescription exposure was associated with deeper LBM loss. In semaglutide-treated patients, corrected mean LBM change worsened from -2.5% in the 1-6 prescription strata to -5.4% in the 7+ prescription strata (p<0.001; **Fig 2E**). In tirzepatide-treated patients, the corresponding worsening was more pronounced, from -1.5% to -7.2% (p<0.001; **Fig 2F**). For patients with obesity and low muscle mass or other features of frailty, these findings suggest that prolonged tirzepatide exposure may warrant caution unless weight-loss goals can reasonably be achieved within 6 months.

### Baseline clinical features stratify LBM response to semaglutide and tirzepatide

Baseline disease features in the semaglutide-treated cohort separated into two broad directions (**Fig. 3A**). Pre-existing diagnoses of hypertension and GERD were significantly positioned on the positive side of the null, indicating patients suffering from these conditions demonstrated less LBM loss. By contrast, pre-existing diagnoses of hypercholesterolemia, abnormal glucose, cervicalgia (−4.1 percentage points; q<0.001), left knee pain (−4.8 pp; q<0.001), and right knee osteoarthritis (−6.2 pp; q<0.001) were significantly associated with greater LBM loss.

**Fig. 3.**
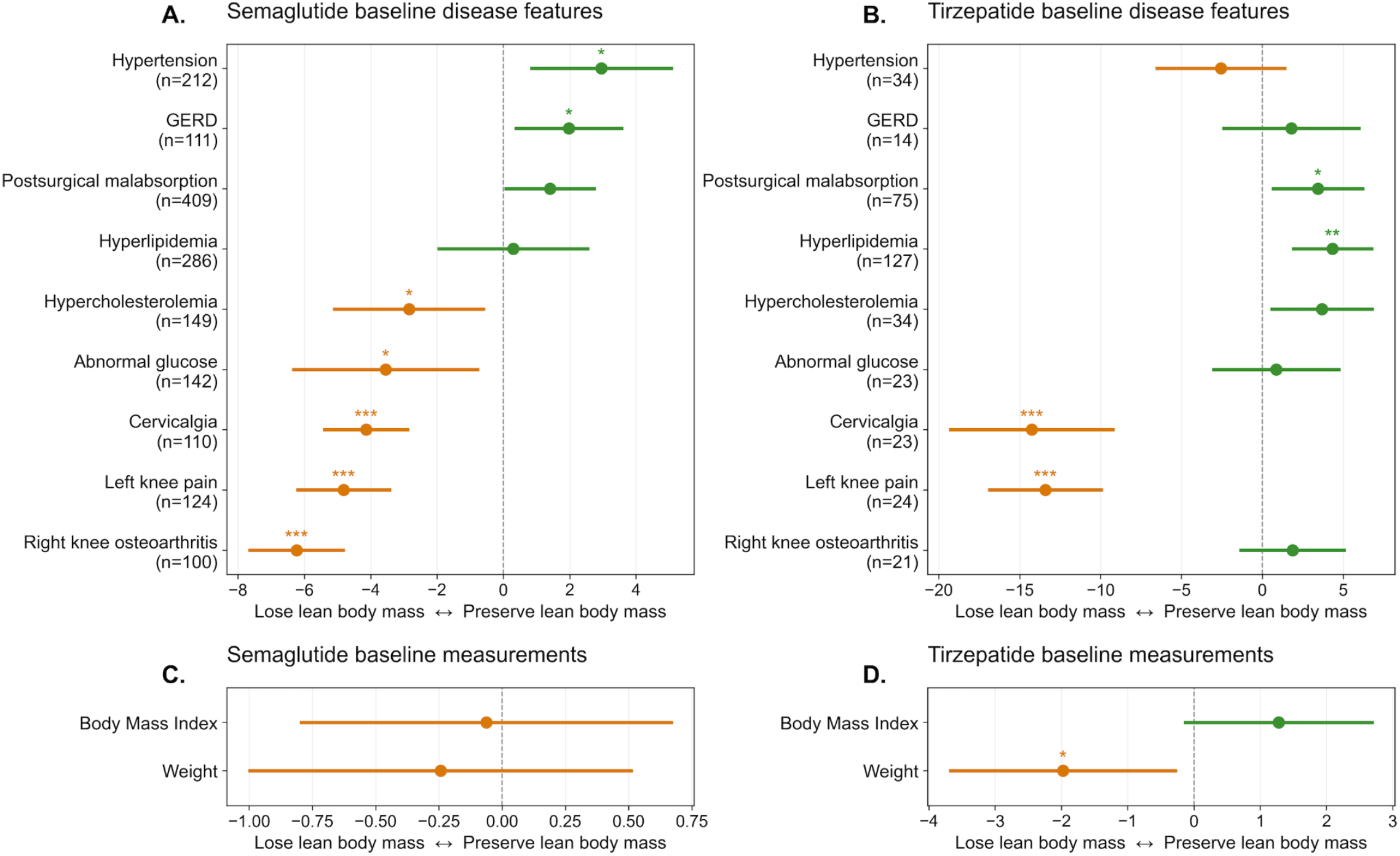
Baseline disease features and measurements associated with lean body mass loss during GLP-1RA treatment. **A**. Pre-existing (baseline) disease feature prevalence in the semaglutide patient cohort presented in Fig. 1 and Fig. 2. Lean body mass loss deviation from the overall semaglutide-treated cohort lean body mass loss response is plotted on the x-axis for the diagnoses presented on the y-axis. Adjusted beta coefficients from multivariable linear regression models are plotted as dots, with horizontal bars indicating 95% confidence intervals. Statistical significance is indicated as * (q<0.05), ** (q<0.01), *** (q<0.001). **B**. As in A, but for the tirzepatide cohort presented in Fig. 1 and Fig. 2. **C**. As in A, but presenting baseline anthropometric measurements (BMI, Weight) for the semaglutide patient cohort presented in Fig. 1 and Fig. 2. For these continuous baseline measures, effect estimates are shown per 1 s.d. increase in the predictor. **D**. As in C, but for the tirzepatide cohort presented in Fig. 1 and Fig. 2.

In the tirzepatide-treated cohort (**Fig. 3B**), the pattern was partly concordant but more heterogeneous than the semaglutide-treated cohort. Hyperlipidemia and postsurgical malabsorption showed the clearest positive associations with less LBM loss. In contrast, cervicalgia (−14.3 pp; q<0.001) and left knee pain (−13.4 pp; q<0.001) again mapped strongly to the negative side, indicating greater LBM loss. Thus, across both agents, pain-associated musculoskeletal features with possible inflammatory elements emerged as the most consistent correlates of greater lean tissue loss. Baseline anthropometric measures showed little discriminatory value in the semaglutide analysis, as both body mass index (BMI) and body weight were centered close to the null and were not associated with abnormal LBM loss compared to the overall semaglutide-treated cohort (**Fig. 3C**). Whereas semaglutide showed near-null associations for both BMI and weight, tirzepatide demonstrated divergence between these measures (**Fig. 3D**): higher BMI was associated with less LBM loss, while higher baseline TBW was associated with greater lean body mass loss. Overall, the data suggest that lean mass response is not explained by body size alone but is instead modified by the broader baseline clinical phenotype. Because several tirzepatide subgroups were markedly smaller than the corresponding semaglutide subgroups, and because the x-axis scales differ between panels, between-drug comparisons should be interpreted as descriptive rather than inferential.

### Greater LBM loss on GLP-1RA may be associated with worse functional outcomes

At 12 months post treatment-initiation, patients with greater LBM loss levels were accompanied by a consistently less favorable functional profile than patients with lower LBM loss levels after both semaglutide treatment (**Fig. 4A**) and tirzepatide treatment (**Fig. 4B**).

**Fig. 4.**
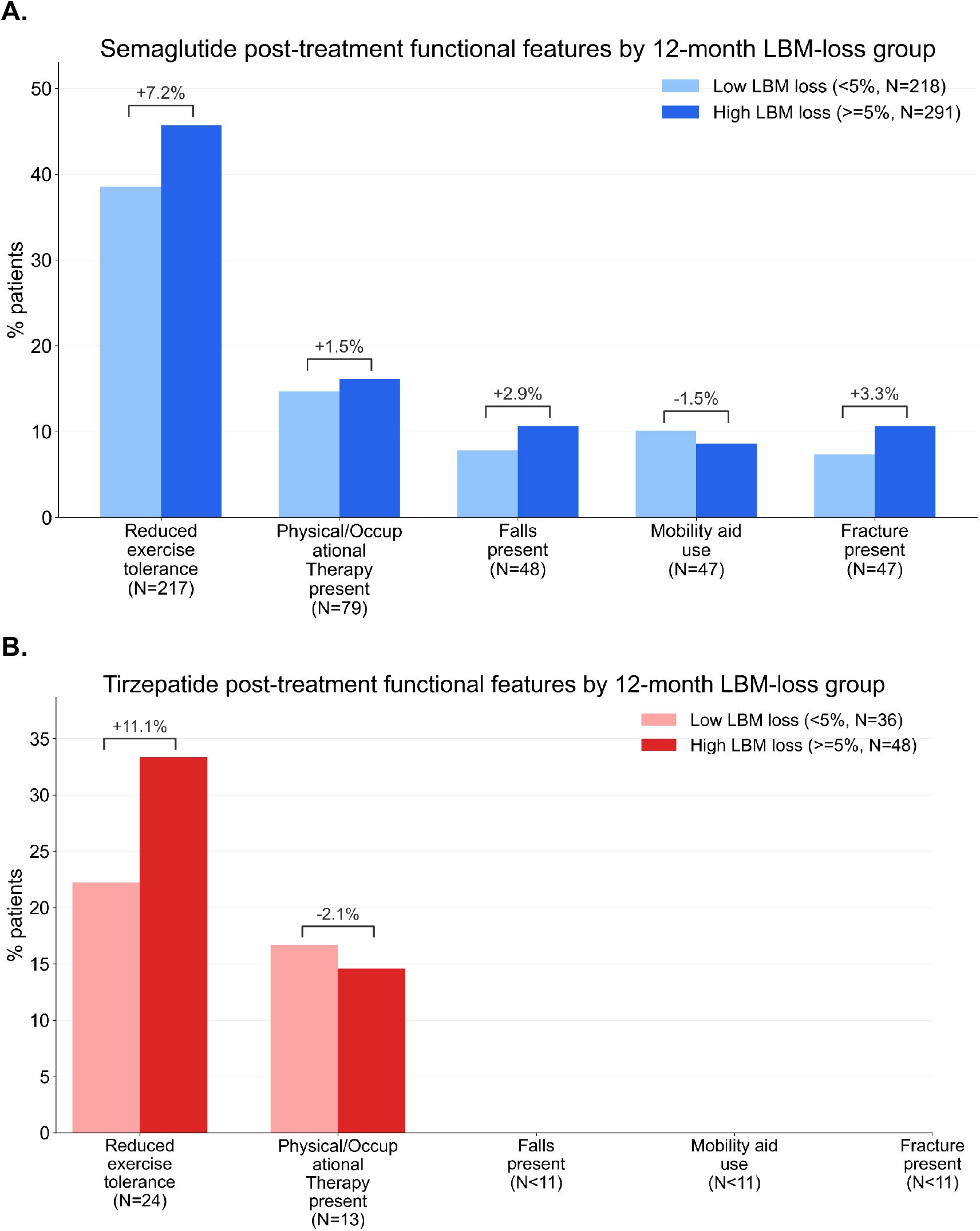
Comparing post-treatment functional outcomes and features for Low and High Lean Body Mass loss subcohorts. **A**. Incidence of patients reporting the following functional outcomes during the 12 months on semaglutide: Reduced exercise tolerance; physical therapy or occupational therapy; any falls; use of a mobility aid; any fractures. Incidence of patients reporting these functional outcomes from the low lean body mass loss group (light blue) are plotted next to the corresponding incidence from the high lean body mass loss group (dark blue) with the delta of inter-group incidence reported above each pair of bars in %age points. **B**. As in A, but for tirzepatide and with the low lean body mass loss group (light red) plotted next to the corresponding high lean body mass group (dark red).

For semaglutide, patients in the high LBM loss group had a higher prevalence of reduced exercise tolerance than those in the low LBM loss group (45.6% versus 38.4%, +7.2 percentage points), and showed modestly higher frequencies of physical or occupational therapy use (16.0% versus 14.5%, +1.5 percentage points), falls (10.7% versus 7.8%, +2.9 percentage points), and fracture presence (10.6% versus 7.3%, +3.3 percentage points) (**Fig. 4A**). Mobility aid use was the only feature that trended in the opposite direction, with a slightly lower prevalence in the high lean body mass loss group (8.5% versus 10.0%, -1.5 percentage points). Natural language processing of clinical notes also found increased mention of ‘fatigue’ in the high LBM loss group (reported for 43.3%, n=553), compared to the low LBM loss group (39.6%, n=762; RR=1.1, X^2^=4.1). Overall, the semaglutide panel suggests that higher LBM loss is clustered with reduced exercise capacity and with a broader pattern of post-treatment functional vulnerability.

A similar but sharper pattern was observed with tirzepatide (**Fig. 4B**). Reduced exercise tolerance showed the largest separation between patients in the high LBM loss group vs patients in the low LBM loss group, occurring in 33.3% of the high LBM loss group versus 22.2% of the low LBM loss group, an absolute difference of +11.1 percentage points. In contrast to semaglutide, physical or occupational therapy use was slightly lower in the high LBM loss group than in the low LBM loss group (14.6% versus 16.7%, -2.1 percentage points). Falls, mobility aid use, and fracture presence were documented in <10 patients within each category, consistent with more recent market approval of tirzepatide, and hence not interpretable at this juncture. As with semaglutide, clinical notes of tirzepatide-treated patients showed increased mention of ‘fatigue’ in the high LBM loss group (24.6%, n=205), compared to the low LBM loss group (19.7%, n=269; RR=1.2, X^2^=7.2).

Taken together, both drugs showed enrichment of reduced exercise tolerance among individuals with greater LBM loss, showing one of the strongest functional correlates of this undesirable outcome of tirzepatide and semaglutide therapy. Because the tirzepatide sample sizes were smaller than those for semaglutide, and because these are descriptive percentages rather than adjusted risk estimates, cross-drug contrasts should be interpreted cautiously. Nonetheless, the signal for reduced exercise tolerance associated with greater LBM loss appeared more prominently with tirzepatide than semaglutide, which may be particularly salient to continue monitoring given the greater absolute LBM loss associated with tirzepatide treatment relative to semaglutide treatment.

### Single cell RNA-seq of musculature shows GIPR+ cells span broader immune, stromal, vascular, and contractile niches than GLP1R+ cells

To place the clinical body-composition divergence into biological context, we analyzed single-cell expression of *GLP1R and GIPR* from the CELLxGENE data portal^25^, leveraging 1556 publicly available human single-cell and single-nucleus RNA sequencing datasets corresponding to 86 million cells from 11,633 human donors. We integrated single-cell musculature annotations from receptor-specific GIPR and GLP1R sheets and retained all non-generic cell types present in either dataset, assigning zero where a receptor-specific entry was absent. Within this harmonized musculature ontology, GIPR-positive cell states extended across immune and hematopoietic, stromal, vascular, and contractile compartments, whereas GLP1R appeared more selective (**Fig. 5A**). Ranking musculature cell types by the number of receptor-expressing cells prioritized broad immune and hematopoietic categories, including leukocyte, mononuclear, lymphoid, myeloid, antigen-presenting, macrophage-lineage, and natural-killer-associated subtypes, with additional contribution from fibroblast, perivascular, smooth-muscle and muscle-lineage backgrounds. This organization suggests that the dominant GIPR-positive cell types are not confined to contractile tissue alone but also include immune-surveillance and tissue-remodeling niches in muscle.

**Fig. 5.**
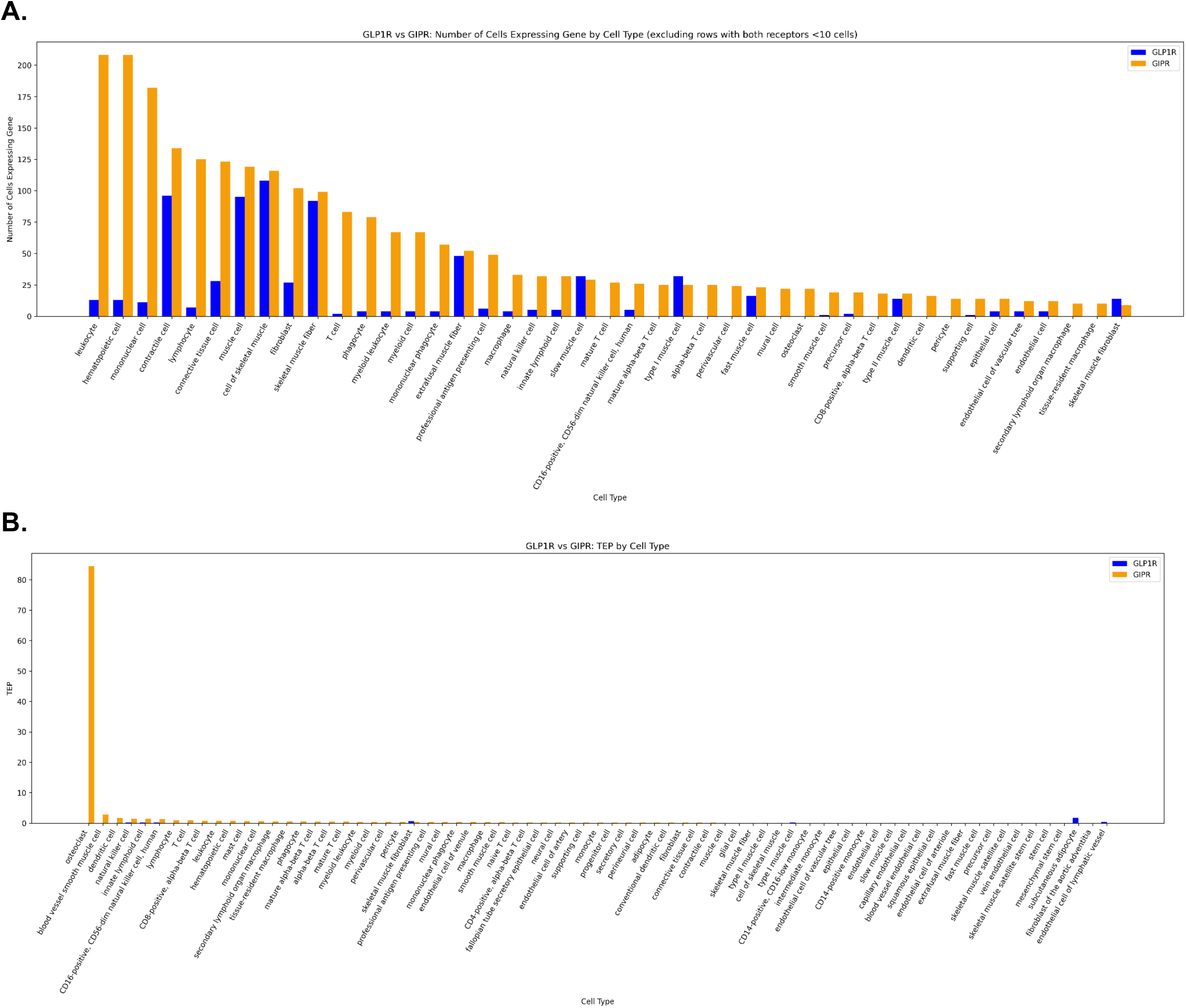
Single-cell RNAseq data demonstrate strength and breadth of GLP1R and GIPR expression in muscle tissue. **A**. GIPR dominates GLP1R in single cell RNA expression across musculature. Number of cells expressing GIPR (orange bars) and GLP1R (blue bars) plotted on the y-axis versus cell types on the x-axis. Cell types with less than 10 individual cells expressing GLP1R and GIPR are not displayed here. Refer to **Fig. S2** in the supplement for the full figure. **B**. The Target Engagement Potential (TEP, see methods) of GLP1R (blue bars) and GIPR (orange bars) are compiled across the musculature tissue from single cell RNA-seq data sets, including all cell types expressing either of these genes significantly. Same Fig. 5B is shown in **Fig. S2** after removing the largest TEP signal for Osteoclast, to enable visualization of the remainder of the moderate TEP signals across muscle tissue.

We next compared target engagement potential (TEP) across matched musculature cell types for GIPR and GLP1R. In the current working analyses, osteoclast emerged as a marked GIPR-enriched outlier, and additional prioritization extended to dendritic, macrophage-related, vascular smooth-muscle and other immune-linked cell states (**Fig. 5B; Fig. S2**). These TEP-based observations are best regarded as hypothesis-generating and do not yet establish a causal mechanism for the clinical body-composition differences.

## Discussion

These findings highlight an important reframing of how GLP-1 based therapies should be evaluated, namely that efficacy cannot be defined by total body weight loss alone but must account for the quality of that weight loss in terms of body composition.^14^. Although tirzepatide delivers superior weight reduction compared with semaglutide, the accompanying increase in lean body mass (LBM) loss (**Figure 1**) suggests that more potent energy deficit induction may come at the cost of greater muscle catabolism, potentially mediated by higher degrees of caloric restriction, altered protein turnover, or differential GIP and GLP-1 signaling effects. Importantly, the weak coupling between total weight loss and LBM loss indicates that these processes are at least partially biologically dissociable, raising the possibility that muscle preserving weight loss is an achievable therapeutic goal rather than an inherent trade off. The small proportion of patients achieving the elite phenotype, defined by high weight loss with minimal LBM loss, further suggests the presence of underlying heterogeneity in treatment response, potentially driven by baseline metabolic state, protein intake, physical activity, or drug exposure patterns. Clinically, even modest LBM losses may have outsized consequences in vulnerable populations, particularly older adults, where sarcopenia and functional decline are closely linked to morbidity. Collectively, these results argue for a paradigm shift toward integrated metabolic phenotyping and support the development of combination strategies, pharmacologic or behavioral, that explicitly target preservation of lean mass alongside adiposity reduction.

Higher dose and cumulative exposure to incretin-based therapies are associated with progressively greater LBM loss (**Figure 2**), extending the earlier observation that weight reduction and muscle loss are only partially coupled. This exposure–response relationship suggests that intensification of therapy amplifies a generalized catabolic state rather than selectively enhancing adipose tissue loss. While greater total body weight reduction with increasing dose and prescription exposure is expected, the marked heterogeneity in LBM trajectories indicates that muscle preservation remains highly variable and is not solely determined by drug intensity. This reinforces the concept that favorable body composition outcomes are achievable but not guaranteed, even at similar exposure levels. The differential patterns observed between semaglutide and tirzepatide further refine this framework, with tirzepatide showing continued weight loss gains at higher exposure but with a steeper trade-off in LBM loss, suggesting diminishing returns from a body composition perspective. Clinically, these data raise important considerations regarding duration and intensity of therapy, particularly in patients with low baseline muscle mass or elevated frailty risk, where prolonged high exposure may shift the risk–benefit balance unfavorably. Taken together, these results support a move toward exposure-aware treatment strategies that integrate longitudinal body composition monitoring, with the goal of achieving metabolic efficacy while minimizing unintended loss of functional tissue.

Semaglutide and tirzepatide produce substantial body-weight reduction in obesity and body-composition analyses indicate that most of the lost tissue is fat mass, although a meaningful absolute reduction in lean mass also occurs^4,26^. Against that background, we find that the baseline phenotype may influence where an individual lies along the spectrum from preferential fat loss to a more mixed fat-and-lean loss response (**Figure 3**). The most reproducible signal across both drugs was a musculoskeletal pain phenotype, especially cervicalgia and knee pain, associated with greater lean body mass loss^27,28^. This is biologically plausible, because chronic pain and mobility limitation can reduce habitual loading and resistance-type activity, thereby aligning with a sarcopenic-obesity phenotype in which preservation of contractile tissue during weight loss is more difficult^12,29,30^. Not all signals were concordant across agents. Hypertension was associated with relative lean-mass sparing in the semaglutide panel but not in the tirzepatide panel; similarly, abnormal glucose and hypercholesterolemia shifted negatively with semaglutide yet positively with tirzepatide. These differences may reflect true biological heterogeneity, but they are also compatible with residual confounders, smaller subgroup sizes and differing model scales. The apparently favorable direction of postsurgical malabsorption is particularly hypothesis-generating and merits external validation. Importantly, lean body mass should not be equated automatically with skeletal muscle mass, since lean tissue estimates can also incorporate body water and non-muscle organs; a reduction in lean mass therefore does not, by itself, establish clinically meaningful sarcopenia^8,31^.

Functional consequences of lean body mass loss are evident in real-world clinical signals following GLP-1 based therapy (**Figure 4**). These data extend the body composition findings by linking higher lean body mass loss to clinically observable changes in patient experience, with reduced exercise tolerance and increased mention of fatigue emerging as the most consistent features across both semaglutide and tirzepatide in patients with greater muscle loss. This suggests that the impact of lean tissue decline is not confined to compositional measurements but is reflected in day-to-day physical capacity and functional status. This interpretation is biologically plausible, as lean mass plays a central role in maintaining strength, mobility, and metabolic resilience, and its loss may compromise physical performance even in the context of overall weight reduction. Although lean mass loss during GLP-1 based therapy is variable across individuals, these findings reinforce the importance of preserving muscle related tissue as a key determinant of clinically meaningful outcomes.

Extending the exposure-dependent and functional consequences of lean body mass loss to underlying biology, the broader cellular distribution of GIPR within musculature provides a plausible mechanistic framework for the differential body composition effects observed clinically. Single cell RNA sequencing reveals that GIPR positive cells span not only contractile muscle lineages but also immune, stromal, and vascular compartments, in contrast to the more restricted expression pattern of GLP1R. This expanded footprint suggests that GIP mediated signaling may influence a wider network of tissue remodeling, immune surveillance, and microvascular processes within muscle, potentially amplifying downstream effects on lean tissue integrity. The enrichment of GIPR in macrophage, dendritic, and osteoclast associated cell states further raises the possibility that immune and inflammatory pathways contribute to muscle turnover during therapy, rather than effects being confined to myocyte metabolism alone. While these observations are hypothesis generating, they offer a mechanistic lens through which the greater lean mass loss seen with dual GIP and GLP 1 receptor agonism can be interpreted and suggest that differences in receptor biology may translate into system level effects on muscle homeostasis.

This study has several important limitations that should be considered when interpreting the findings. First, the observational design precludes causal inference, and despite large-scale real-world data and adjustment strategies, residual confounding related to treatment selection, baseline health status, lifestyle factors, and unmeasured clinical variables is likely. Second, body composition measurements were derived from heterogeneous modalities, including BIA, DEXA, and air displacement plethysmography, each with differing precision and susceptibility to measurement variability, which may introduce noise into longitudinal LBM estimates. Third, drug exposure was inferred from prescription patterns rather than confirmed adherence, and out-of-system medication use, including compounded or cash-pay GLP-1 therapies, may not be fully captured. Fourth, functional outcomes such as fatigue and exercise tolerance were derived from clinical documentation and are subject to variability in reporting, clinician interpretation, and natural language processing extraction. Fifth, the single cell analyses are hypothesis generating and do not establish causal mechanisms, as receptor expression does not directly equate to functional signaling or pharmacologic effect in vivo. Finally, the subset of patients with paired body composition data represents a small fraction of the overall treated population, raising the possibility of selection bias toward individuals undergoing more intensive monitoring. Collectively, these limitations underscore the need for prospective studies integrating standardized body composition assessment, objective functional endpoints, and mechanistic validation to more definitively characterize the relationship between incretin therapy, muscle biology, and clinical outcomes.

In summary, GLP-1 based therapies achieve substantial weight loss but are accompanied by variable and, in some patients, clinically meaningful reductions in lean body mass that scale with drug exposure. The dissociation between total weight loss and muscle preservation, together with the emergence of functional signals such as fatigue and reduced exercise tolerance, underscores the importance of shifting from weight-centric to composition-aware therapeutic goals. The broader biological footprint of dual GIP and GLP-1 receptor agonism further suggests that differences in receptor biology may contribute to these effects. Taken together, these findings highlight an unmet need for strategies that preserve lean mass while maintaining metabolic efficacy and support the integration of body composition monitoring into both clinical practice and future therapeutic development.

## Methods

### Data Source and Study Design

We conducted a retrospective cohort study using deidentified longitudinal electronic health record data from the nSights Federated EHR Network^5–7,33^. All analyses were conducted on deidentified data. The study was designed to compare longitudinal body-composition changes after initiation of semaglutide or tirzepatide, characterize the relationship between lean body mass (LBM) loss and weight loss, evaluate dose-response patterns, identify baseline features associated with subsequent LBM loss, and assess post-treatment functional correlates of LBM loss.

### Study Cohorts and Exposure Definition

We identified adults with at least one prescription for semaglutide or tirzepatide. For the primary body-composition analyses, we retained a single treatment episode per patient, defined as the first observed GLP-1 receptor agonist (GLP-1RA) episode for either semaglutide or tirzepatide. Patients were assigned to the exposure group according to the drug initiated at that first GLP-1RA episode, regardless of any subsequent switching. The episode start date was defined as the date of the first prescription for the index drug, and the episode end date was defined as the last observed prescription date for that episode.

Across 670,422 first-episode GLP-1 patients, 456,742 initiated semaglutide and 213,680 initiated tirzepatide (**Fig. 1A**). Of these, 7,965 patients had baseline LBM measurements before GLP-1RA initiation and follow-up LBM measurements after treatment initiation, including 6,196 semaglutide-treated and 1,769 tirzepatide-treated individuals. These patients constituted the primary analytic cohort for the body-composition analyses presented in this manuscript.

For dose-response analyses, exposure was defined using the maximum achieved dose within the first 12 months after treatment initiation. Dose bands were based on clinically used dose levels for semaglutide and tirzepatide. We additionally summarized treatment intensity using the number of prescriptions recorded during follow-up, which was used in descriptive dose-by-prescription heatmaps. Treatment duration was derived from longitudinal prescribing records and summarized descriptively.

### Large Language Model-Based Body Composition Extraction and Measurement Ascertainment

To identify body-composition measurements, we first enriched the corpus of clinical notes using a broad keyword strategy targeting body-composition language, including lean body mass, fat mass, fat percentage, muscle mass, fat-free mass, Tanita, and related terminology. We then applied a note-level large language model (LLM) extraction workflow to those candidate notes. The extraction model was gpt-oss-20b with structured JSON output. Each prompt included the full note text together with the structured note date, and the model was instructed to extract measurement date, measurement source, body weight, fat percentage, lean body mass and supporting evidence text from each note (see **Supplementary Methods**).

Extraction outputs were further harmonized. Units were standardized, with lean body mass and fat mass converted to kilograms and weight measurements converted to kilograms when required. To resolve unit inconsistencies, we used explicit unit strings in the extracted output, unit cues in the note evidence text, parenthetical metric equivalents embedded in the note (for example, “132 lb (59.9 kg)”), arithmetic consistency among total body weight, lean body mass, and fat percentage, and proximal structured weight measurements when available. Fat mass was derived when necessary, from total body weight and fat percentage, and muscle percentage was defined as “100 − fat percentage”.

Measurement sources extracted by the LLM were canonicalized into a small set of device-level categories, including Tanita, generic bioimpedance, Seca, DEXA, Bod Pod, indirect calorimetry, scale, and other. Structured body weight and body mass index (BMI) were obtained from structured measurement tables.

### Baseline Covariates

Baseline covariates included age at index, sex as recorded in the EHR, baseline BMI, baseline structured weight, baseline LBM, baseline fat mass, baseline muscle percentage, prior GLP-1RA exposure, and baseline type 2 diabetes mellitus (T2DM) status. Baseline LBM and weight were defined as the value closest to treatment initiation within a window from 180 days before to 14 days after the index date. Baseline BMI was defined as the closest structured BMI value from 365 days before through 14 days after the index date. Prior GLP-1RA exposure was defined by any pre-index exposure to another GLP-1RA agent, including agents such as liraglutide or dulaglutide. Baseline T2DM was defined using ICD-10 E11* codes and ICD-9 type 2 diabetes equivalents, requiring at least three diagnosis dates before treatment initiation

### Longitudinal Body-Composition Analyses

For the primary longitudinal analyses (**Fig. 1B**), we evaluated changes in LBM and total body weight at 3, 6, 9, and 12 months after treatment initiation. Fixed landmark windows were used: 3, 6, and 9 months were evaluated using ±30-day windows, and 12 months was evaluated using a ±45-day window. Within each time window, if multiple values were available for a patient, the patient-level median was used. LBM and total body weight were analyzed both as percent change from baseline, calculated as 100 × (follow-up − baseline) / baseline, and as absolute change from baseline in kilograms when applicable. Trajectory plots display means with standard error bars.

### Dose-Response Analyses

For dose-response analyses, we paired each patient’s maximum achieved dose within 12 months after treatment initiation with that patient’s maximum achieved change in lean body mass (**Fig. 2A-B**) or total body weight (**Fig. 2C-D**) during the same 12-month period. These figures display dose-binned summary estimates rather than individual patient-level regression fits. Dose categories corresponded to labeled dosing increments for injectable semaglutide formulations (Ozempic and Wegovy): 0.25 mg, 0.5 mg, 1.0 mg, 1.7 mg, 2.0 mg, and 2.4 mg or greater, and for tirzepatide (Mounjaro and Zepbound): 2.5 mg, 5.0 mg, 7.5 mg, 10.0 mg, 12.5 mg, and 15.0 mg. Observed dose values were used when directly available in the medication record, and missing dose values were extracted from medication descriptions using an LLM-assisted dose-mapping workflow^34^. To reduce the influence of extreme values, LBM values only were winsorized within each dose group using Tukey fences (Q1-1.5xIQR and Q3+1.5xIQR) before summary estimation and fitting; TBW values were not winsorized. A linear regression was fitted to the dose-level mean LBM change percentage to quantify the overall trend across dose categories; where reported, R^2^ reflects fit across these dose-level summary means rather than variance explained at the individual-patient level.

For the prescription count by dosage heatmaps (**Fig. 2E-H**), we classified each patient according to the maximum achieved dose within the first 12 months after treatment initiation and the total number of GLP-1RA prescriptions recorded during that same 12-month period, grouped as 1-3, 4-6, 7-9, and 10 or more prescriptions. Heatmaps were generated separately for LBM change and TBW change. To ensure direct comparability across outcomes, the TBW heatmaps were restricted to the same analytic subcohort of patients contributing LBM measurements. Within each dose-prescription cell, LBM was summarized using the same Tukey-fence winsorization approach, whereas TBW was summarized without winsorization. For cells containing fewer than 11 patients, the mean value was suppressed and displayed as N/A in the heatmaps.

For dose-exposure modeling, we analyzed LBM change and total body weight change using separate bin-level linear models for semaglutide and tirzepatide, with dosage and prescription-count stratum modeled as categorical predictors together with a dose-by-exposure interaction term. This approach was used because neither dose nor treatment exposure was assumed to have a strictly linear effect across strata. Statistical significance was determined with F-tests comparing models with and without the terms of interest. Specifically, the overall joint association of dose and treatment exposure was assessed by comparing the full dose-exposure model with an intercept-only model, whereas the independent contribution of dose or prescription exposure was assessed by comparing models that retained the other predictor. Where reported, differences between prescription-count strata were estimated within the same model after including dose-band terms, so that those p-values reflect exposure-associated differences after accounting for dose.

### Baseline Feature Analyses

We next evaluated associations between baseline clinical features and subsequent LBM loss (**Fig. 3**). The primary outcome for these analyses was the highest observed 12-month LBM percent change after treatment initiation. Baseline disease-related features were defined from diagnosis data in the two years before treatment initiation and required at least three occurrences on distinct dates to be considered present. A total of 3,746 unique ICD-coded disease features were evaluated across both drugs (3,308 for semaglutide and 1,730 for tirzepatide, with 1,292 features common to both cohorts). For each drug separately, we fit multivariable linear regression models with heteroscedasticity-robust standard errors, modeling 12-month LBM percent change as a continuous outcome. Disease-feature models included the candidate baseline disease feature together with age at index, sex, baseline BMI, baseline weight, index year, and baseline type 2 diabetes status as covariates. Quantitative baseline-feature models used the same adjustment set with continuous standardized predictors. Multiple testing across baseline disease features was controlled within drug using the Benjamini-Hochberg (BH) false discovery rate procedure. Figure 3 displays regression coefficients with 95% confidence intervals for baseline disease and measurement features retained for plotting; disease features were required to have at least 100 affected patients for a drug.

### Functional Outcome Analyses

Finally, we examined whether greater LBM loss was associated with post-treatment functional features (**Fig. 4**). Patients were classified into low- and high-LBM-loss groups on the basis of 12-month LBM change, with high loss defined as at least 5% loss from baseline and low loss defined as less than 5% loss. Five binary functional features were derived from post-treatment clinical documentation using an LLM-based extraction workflow (similar to body composition extraction) and included reduced exercise tolerance, physical or occupational therapy use, falls, mobility-aid use, and fracture (see **Supplementary Methods**). For each drug, we summarized the prevalence of these features in the high- and low-LBM-loss groups. The figure displays group-specific prevalence and absolute percentage-point differences between groups.

### Analysis of GLP1R and GIPR Expression Using Single-Cell

Single-cell expression of *GLP1R and GIPR* was obtained from the CELLxGENE data portal^16^ (https://cellxgene.cziscience.com/), leveraging 1556 publicly available human single-cell and single-nucleus RNA sequencing datasets corresponding to 86 million cells from 11,633 human donors. Using the CELLxGENE Census, GLP1R and GIPR expression was obtained across available cell types. Gene expression values were extracted as normalized counts (counts per 10,000 [CP10K], log-transformed where applicable) and analyzed at the cell-type level using curated annotations provided within each dataset. For each cell type, *GLP1R and GIPR* expression was summarized as both mean normalized expression and the proportion of expressing cells (non-zero counts). Where multiple datasets contributed to the same tissue, results were aggregated to ensure robustness across cohorts, and analyses were restricted to cell types with adequate representation to minimize sparsity-driven artifacts.

Single-cell GLP1R atlas data were analyzed as follows. For each tissue-cell annotation, Expression (CP10K), number of GLP1R-positive / GIPR-positive cells, and percentage of expressing cells were analyzed.

The primary metric was *Target Engagement Potential (TEP)*, defined as Expression (CP10K) × (% expressing cells / 100). Conceptually, TEP is a prevalence-weighted expression metric: it becomes high when a cell class has both appreciable target receptor transcript abundance and a meaningful proportion of cells that are target-positive. It is therefore useful for ranking which cellular compartments are most likely to be engaged under a broad systemic exposure model because it balances intensity and prevalence rather than privileging either one alone. A compartment with very high expression in only a tiny number of cells may not rank as highly by TEP as a compartment with slightly lower expression but a much broader target-positive fraction.

### Statistical Analysis

Continuous variables are presented as mean (standard deviation) or median [interquartile range], as appropriate, and categorical variables are presented as counts and percentages. Longitudinal body-composition summaries were generated from patient-level values within each prespecified time window and then summarized within each treatment group. Trajectory plots display group means with standard error bars. Intergroup comparisons shown on the trajectory plots were based on approximate two-sample Welch-type tests. Comparisons of categorical distributions across treatment groups, including heatmap-derived subgroup comparisons, were performed using Pearson chi-square tests, with Fisher exact tests used where appropriate. Overall significance for the dose-exposure analyses was assessed with F-tests comparing the full model against an intercept-only model. The independent contributions of dose and prescription exposure were evaluated with nested-model F-tests, and dose-adjusted comparisons between prescription-count strata were estimated within the same modeling framework.

For baseline feature analyses, multivariable linear regression was used to model 12-month lean body mass percent change as a continuous outcome. Models of baseline disease features adjusted for age at index, sex, baseline BMI, baseline weight, index year, and baseline type 2 diabetes status. Baseline measurement features were analyzed using analogous multivariable linear models with continuous standardized predictors. Multiple testing across baseline disease features was controlled using the Benjamini-Hochberg false discovery rate procedure. For the functional analyses, results are reported as effect estimates / prevalence differences.

### Data Source

This study analyzed de-identified EHR data from academic medical centers in the United States via the nference nSights Analytics Platform. Prior to analysis, all data underwent expert determination de-identification satisfying HIPAA Privacy Rule requirements (45 CFR §164.514(b)(1)), employing a multi-layered transformation approach for both structured data (cryptographic hashing of identifiers, date-shifting, geographic truncation) and unstructured clinical text (ensemble deep learning and rule-based methods with >99% recall for personally identifiable information detection)^35,36^. nference established secure data environments within each participating center, housing these de-identified patient data governed by expert determination. These de-identified data environments were specifically designed to enable data access and analysis without requiring Institutional Review Board oversight, approval, or exemption confirmation. Accordingly, informed consent and IRB review were not required for this study.

### Data Availability

This study involves the analysis of de-identified Electronic Health Record (EHR) data via the nference nSights Federated Clinical Analytics Platform (nSights). Data shown and reported in this manuscript were extracted from this environment using an established protocol for data extraction, aimed at preserving patient privacy. The data has been de-identified pursuant to an expert determination in accordance with the HIPAA Privacy Rule. Any data beyond what is reported in the manuscript, including but not limited to the raw EHR data, cannot be shared or released due to the parameters of the expert determination to maintain the data de-identification. The corresponding author should be contacted for additional details regarding nSights.

### De-identification and HIPAA compliance certification

Prior to analysis, all EHR data were de-identified under an expert determination consistent with the Health Insurance Portability and Accountability Act (HIPAA) Privacy Rule (45 CFR §164.514(b)(1)). The de-identification methodology employed a multi-layered transformation approach to both structured and unstructured data fields^35,36^. In structured data, direct identifiers including patient names and precise geographic locations were excluded entirely, while indirect identifiers underwent specific transformations: patient identifiers, medical record numbers, and accession numbers were replaced with one-way cryptographic hashes using confidential salts to preserve linkage across patient encounters; all dates were shifted backward by patient-specific random offsets (1–31 days) to preserve temporal relationships while obscuring exact event timing; the ZIP codes were truncated to two-digit state-level resolution; and continuous variables including age, height, weight, and body mass index were thresholded to prevent identification of extreme values (for example, ages ≥89 years transformed to ‘89+’ and BMI >40 transformed to ‘40+’). In unstructured clinical text, an ensemble de-identification system that combines attention-based deep learning models with rule-based methods achieved an estimated >99% recall for personally identifiable information (PII) detection, with detected identifiers replaced by plausible fictional surrogates^35^.

### Data Harmonization

To address heterogeneity in EHR data, we harmonized clinical variables including medications, anthropometric measurements, and diagnoses to standardized concepts. For medications, we first constructed a standardized drug concept database combining the nSights knowledge graph with RXNorm (https://www.nlm.nih.gov/research/umls/rxnorm/index.html) hierarchies to capture ingredient, brand, and dose-specific information^37^. EHR medication records were matched using a hierarchical approach prioritizing RXNorm codes when available, followed by ingredient-level matching, and finally natural language processing and pattern matching on free-text medication orders when structured codes were absent. For anthropometric measurements (height, weight, BMI), we created a unified vocabulary from SNOMED (https://www.snomed.org/, https://athena.ohdsi.org) and LOINC (https://loinc.org/) terminologies and matched EHR measurement descriptions using standardized text matching algorithms with abbreviation expansion and synonym resolution; ambiguous mappings were resolved using OpenAI GPT-4o (https://platform.openai.com/docs/models/gpt-4o) with summary statistics as context, followed by manual verification. For diagnoses, we developed a hierarchical disease concept database from the nSights knowledge graph and matched EHR diagnosis descriptions and codes by identifying the most specific common child concept in the hierarchy. This approach enabled consistent identification of clinical entities while preserving granularity where available.

## Conflict of Interest Statement

The authors are employees of nference, inc., which conducts research collaborations with various biopharmaceutical companies whose therapeutic products are included in this study. None of these companies, nor any other nference collaborator, funded, supported, or had any role in the independent study design, data acquisition, analysis, interpretation, manuscript preparation, or the decision to submit this work for publication. All analyses were conducted by the authors using de-identified electronic health record data. The authors declare no additional competing interests.

## Acknowledgements

We thank the nference engineering team for the development of the nSights federated AI platform, and Patrick Lenehan and Christopher Gregg for critical review and feedback.

## SUPPLEMENTARY MATERIAL

**Fig. S1.**
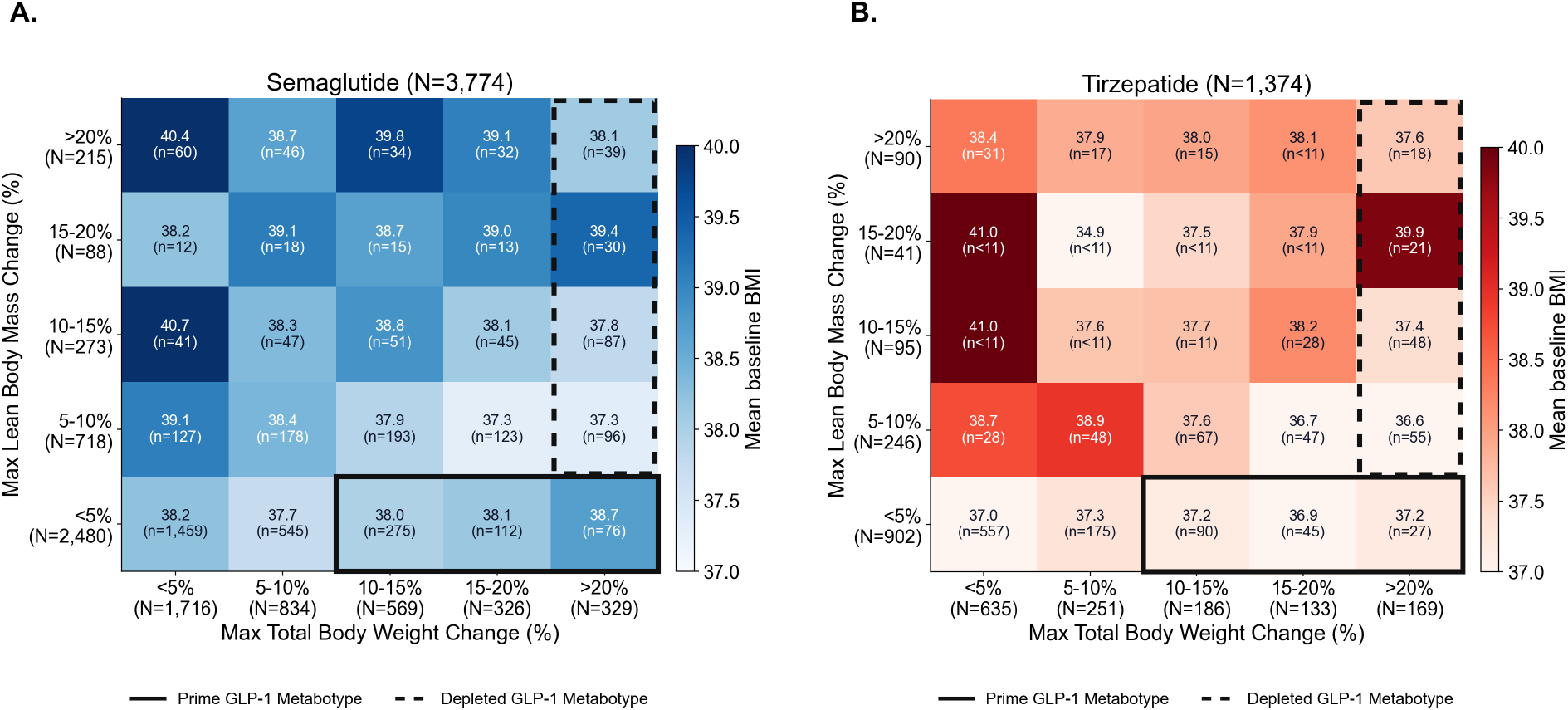
Mean baseline BMI across GLP-1 receptor agonist response strata for semaglutide and tirzepatide. Heatmap of mean baseline BMI across patient response strata defined by maximum % total body weight change (x-axis: <5%, 5–10%, 10–15%, 15–20%, >20% loss) and maximum % lean body mass change (y-axis: <5%, 5–10%, 10–15%, 15–20%, >20% loss) for **(A)** semaglutide (N=3,774) and **(B)** tirzepatide (N=1,374). Each tile displays the mean baseline BMI and patient count. Tile color intensity reflects mean BMI magnitude. The solid box denotes the *Prime GLP-1 Metabotype* (≥10% total body weight loss with <5% lean body mass loss), and the dashed box denotes the *Depletive GLP-1 Metabotype* (>20% total body weight loss with ≥5% lean body mass loss).

**Fig. S2.**
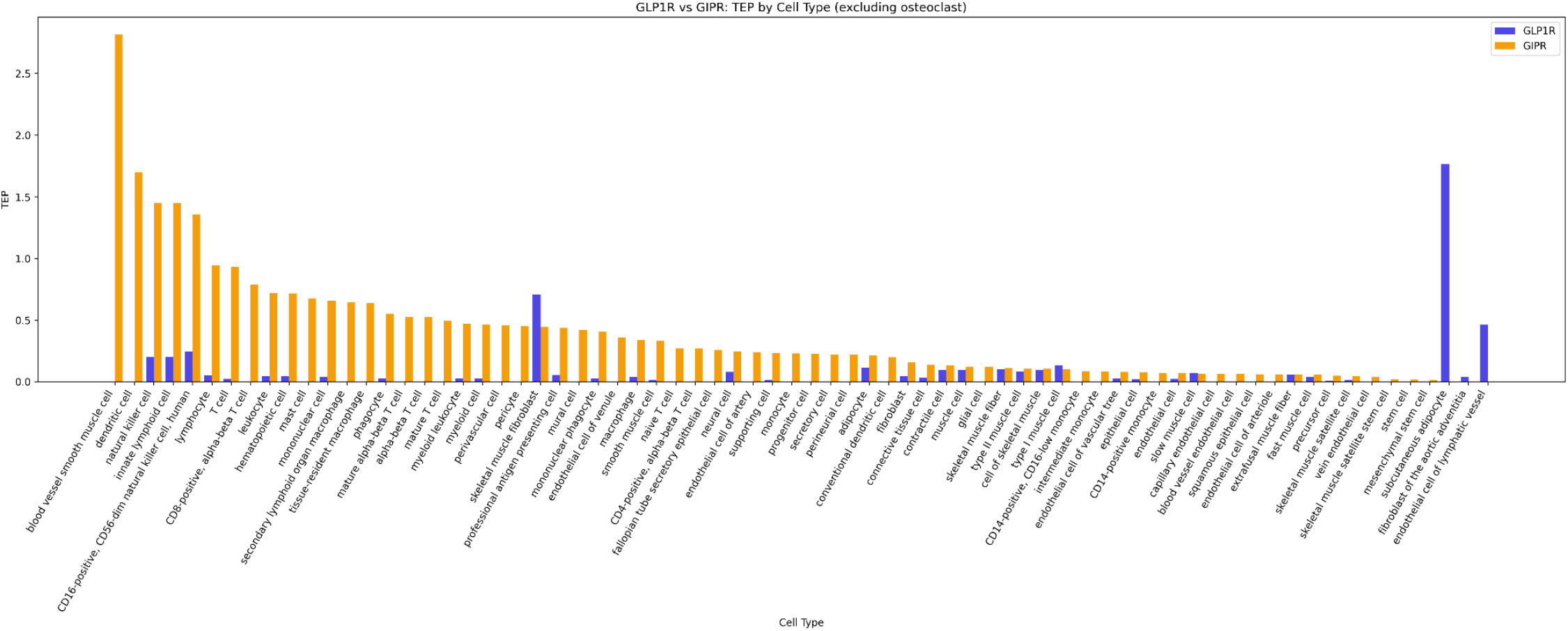
The Target Engagement Potential (TEP, see methods) of GIPR (orange bars) and GLP1R (blue bars) are compiled across the musculature tissue from single cell RNA-seq data sets, including all cell types expressing either of these genes significantly.

## Supplementary Methods

### Large Language Model-Based Extraction of Body Composition and Functional Outcomes

Clinical note extraction was performed using GPT-OSS-20B on NVIDIA A100 40GB GPUs. For both body-composition and functional-outcome extraction, model generation used a temperature of 0.6 and a maximum output length of 8,192 tokens.

#### Body composition extraction

Candidate notes for body-composition extraction were identified using the following exact screening terms:

- fat %
- body fat percent
- percent body fat
- % fat
- lean body mass
- muscle mass
- fat mass
- basal metabolic rate
- bmr
- tanita
- fat free mass System prompt

#### System prompt

You are a clinical information extraction system. Extract body composition measurements from clinical notes and return valid JSON only.

#### User prompt

Note: “<YYYY-MM-DD>“ and “<note text>“ are replaced with structured note date and full note text respectively, at inference time

Extract body-composition measurements from this clinical note.

<<<NOTE_FRAGMENT_START>>>

Structured note date: <YYYY-MM-DD>

Clinical note: <note text>

<<<NOTE_FRAGMENT_END>>>

Use the structured note date as the temporal anchor. If the note clearly refers to a prior or future measurement, attribute the measurement to that other time as best as possible. Do not assume every value is from the note date. If a precise date is unavailable, use the best approximate temporal information present in the note.

Return ONLY a JSON array. Each item in the array must represent one measurement timepoint/event and use this schema:

[

{

“measurement_date”: “YYYY-MM-DD or null”,

“measurement_date_text”: “verbatim or normalized temporal phrase”,

“date_precision”: “exact|month|year|relative|note_date|unknown”,

“date_relation_to_note”: “same_day|before_note|after_note|unknown”,

“measurement_source”: “canonical lowercase comma-separated source list or null”,

“body_weight”: {“value”: number, “unit”: string} or null,

“fat_percent”: {“value”: number, “unit”: “%”} or null,

“lean_body_mass”: {“value”: number, “unit”: string} or null,

“basal_metabolic_rate”: {“value”: number, “unit”: “kcal”} or null,

“evidence_text”: “short supporting excerpt”

}

]

Rules:

- Include only measurements actually stated or strongly implied in the note.
- Extract only measurements that are actually attributed to the patient, whether current or historical.
- Do not extract values that are suggestions, goals, targets, recommendations, educational examples, template text, or hypothetical/future-only values.
- Do not extract changes, differences, gains, losses, or percentages of change in these metrics; extract only absolute measurement values. For example, “15% decrease in lean mass” should not be extracted as a lean_body_mass value.
- If multiple metrics belong to the same timepoint, place them in the same JSON object.
- If the note contains multiple distinct timepoints, return multiple JSON objects.
- If the note states the measurement source or device, extract it as measurement_source using canonical lowercase labels.
- If more than one source applies to the same timepoint, return a comma-separated lowercase list such as “tanita, scale”.
- Obvious examples include: “dexa”, “tanita”, “inbody”, “bod pod”, “bioimpedance”, “skinfold”, “scale”. Use null if the source is not stated.
- Interpret “ffm” and “fat free mass” as lean body mass when the note is using them as equivalent body-composition metrics.
- Preserve the units stated in the note. Do not convert units.
- Prefer “kg” for body_weight and lean_body_mass when the note provides kilograms explicitly.
- For fat_percent, always use unit “%”.
- For basal_metabolic_rate, always use unit “kcal”.
- If none of Basal Metabolic Rate, Fat %, Lean Body Mass, or Body weight are present, return [].
- Output must be valid JSON with no markdown fences and no extra commentary.

#### Example structured output

[

{

“measurement_date”: “2024-09-22”,

“measurement_date_text”: “current visit”,

“date_precision”: “exact”,

“date_relation_to_note”: “same_day”,

“measurement_source”: “tanita”,

“body_weight”: {“value”: 78.7, “unit”: “kg”},

“fat_percent”: {“value”: 39.8, “unit”: “%”},

“lean_body_mass”: {“value”: 47.4, “unit”: “kg”},

“basal_metabolic_rate”: {“value”: 1441, “unit”: “kcal”},

“evidence_text”: “Tanita assessment performed at the current visit showed weight 78.7 kg, body fat 39.8%, lean body mass 47.4 kg, and basal metabolic rate 1441 kcal.”

}

]

##### 2. Functional outcome extraction

Candidate notes for functional-outcome extraction were identified using the following exact screening terms:

- fall
- falls
- fracture
- physical therapy
- occupational therapy
- pt evaluation
- ot evaluation
- walker
- rollator
- cane
- wheelchair
- crutches
- mobility aid
- assistive device
- weakness
- decondition
- frail
- exercise tolerance
- endurance
- functional status

#### System prompt

You are a clinical information extraction system. Extract functional-status and physical-performance data from clinical notes and return valid JSON only.

#### User prompt

Note: “<YYYY-MM-DD>“ and “<note text>“ are replaced with structured note date and full note text respectively, at inference time

Extract functional metrics from this clinical note.

<<<NOTE_FRAGMENT_START>>>

Structured note date: <YYYY-MM-DD>

Clinical note: <note text>

<<<NOTE_FRAGMENT_END>>>

Use the structured treatment start date and structured note date as temporal anchors. If the note clearly refers to a prior or future functional assessment, attribute that assessment to the best available date. Do not assume every metric occurred on the note date. If a precise date is unavailable, use the best approximate temporal information present in the note.

Return ONLY a JSON array. Each item in the array must represent one functional-assessment event/timepoint and use this schema:

[

{

“assessment_date”: “YYYY-MM-DD or null”,

“assessment_date_text”: “verbatim or normalized temporal phrase”,

“date_precision”: “exact|month|year|relative|note_date|unknown”,

“date_relation_to_note”: “same_day|before_note|after_note|unknown”,

“falls”: {“status”: “present|absent|unknown”, “count”: number or null, “timeframe_text”: string or null} or null,

“fracture”: {“status”: “present|absent|unknown”, “site”: string or null, “timeframe_text”: string or null} or null,

“pt_ot_assessment”: {“status”: “present|absent|unknown”, “discipline”: “pt|ot|pt,ot|unknown”, “summary”: string or null} or null,

“mobility_aid_use”: {“status”: “present|absent|unknown”, “device”: string or null} or null,

“weakness_signal”: {“status”: “present|absent|unknown”, “summary”: string or null} or null, “exercise_tolerance_signal”: {“status”: “reduced|normal|improved|unknown”, “summary”: string or null} or null,

“evidence_text”: “short supporting excerpt”

}

]

Rules:

- Extract only patient-attributed functional information, whether current or historical.
- Include qualitative findings when clearly documented.
- Do not extract goals, recommendations, hypothetical plans, educational examples, template placeholders, or unrelated family history.
- If multiple metrics belong to the same assessment timepoint, place them in the same JSON object.
- If a note contains multiple distinct functional-assessment timepoints, return multiple JSON objects.
- Use “unknown” for qualitative statuses only when the note mentions the domain but does not make the polarity clear.
- For falls and fractures, mark “present” only when the patient is described as having a history/event; mark “absent” only when the note explicitly denies them.
- For PT/OT, extract formal therapy or therapist assessments/evaluations only.
- For mobility aid use, include devices such as cane, walker, wheelchair, rollator, crutches, prosthesis, or other assistive ambulation aids.
- For weakness_signal, capture clinician-described weakness, frailty, deconditioning, sarcopenia-like weakness, or similar functional weakness signals.
- For exercise_tolerance_signal, use “reduced” for poor/decreased/limited exercise tolerance, exertional limitation, poor endurance, or low stamina; use “normal” or “improved” only when the note clearly states that.
- If none of the requested metrics/signals are present, return [].
- Output must be valid JSON with no markdown fences and no extra commentary.

#### Example structured output

[

{

“assessment_date”: “2024-06-10”,

“assessment_date_text”: “today”,

“date_precision”: “note_date”,

“date_relation_to_note”: “same_day”,

“falls”: {“status”: “absent”, “count”: null, “timeframe_text”: null},

“fracture”: null,

“pt_ot_assessment”: {“status”: “present”, “discipline”: “pt”, “summary”: “physical therapy evaluation completed”},

“mobility_aid_use”: {“status”: “present”, “device”: “cane”},

“weakness_signal”: {“status”: “present”, “summary”: “generalized weakness with deconditioning”},

“exercise_tolerance_signal”: {“status”: “reduced”, “summary”: “poor exercise tolerance and limited endurance”},

“evidence_text”: “Physical therapy evaluation completed today. Patient ambulates with a cane, reports generalized weakness and deconditioning, and has poor exercise tolerance with limited endurance. Denies recent falls.”

}

]

##### 3. General considerations

For both extraction tasks, prompts required valid JSON output only and explicitly instructed the model not to extract template text, hypothetical recommendations, educational examples, or values not attributable to the patient. Temporal anchoring language was included to distinguish current, historical, and future-referenced observations. Supporting evidence excerpts were retained with each extracted record to facilitate manual review and downstream quality assurance.

## Notes

### Funding Statement

This study did not receive any external funding.

### Author Declarations

This study analyzed de-identified EHR data from academic medical centers in the United States via the nference nSights Analytics Platform. Prior to analysis, all data underwent expert determination de-identification satisfying HIPAA Privacy Rule requirements (45 CFR 164.514(b)(1)), employing a multi-layered transformation approach for both structured data (cryptographic hashing of identifiers, date-shifting, geographic truncation) and unstructured clinical text (ensemble deep learning and rule-based methods with <99% recall for personally identifiable information detection)35,36. nference established secure data environments within each participating center, housing these de-identified patient data governed by expert determination. These de-identified data environments were specifically designed to enable data access and analysis without requiring Institutional Review Board oversight, approval, or exemption confirmation. Accordingly, informed consent and IRB review were not required for this study. 35.Murugadoss, K. et al. Building a best-in-class automated de-identification tool for electronic health records through ensemble learning. Patterns (N Y) 2, 100255 (2021). 36.Murugadoss, K. et al. Scaling text de-identification using locally augmented ensembles. medRxiv 2024.06.20.24308896 (2024) doi:10.1101/2024.06.20.24308896.

